# Delayed Access to Innovative Medicines in Romania: A Comprehensive Analysis of the Reimbursement Processes (2015-2024)

**DOI:** 10.1101/2025.02.13.25322197

**Authors:** Constantin Radu, Diana Elena Serban, Nona Delia Chiriac

## Abstract

**Introduction:** Romania’s reimbursement framework for innovative medicines relies on health technology assessments (HTAs) resulting in unconditional or conditional decisions. Although conditional decisions aim to manage financial uncertainty via Cost-Volume (CV), anecdotal evidence points to growing delays and a growing backlog of indications waiting to be reimbursed. This study is the first to systematically quantify these delays and assess their evolution over time.

**Methods:** We analyzed all publicly available full HTA reports (2015–2024) from Romania’s National Drug Agency. Each indication was classified by HTA decision (unconditional, conditional, or negative) and reimbursement status. Descriptive analyses included mean and median durations for HTA and reimbursement processes. A Kaplan-Meier survival analysis compared time-to-reimbursement between conditional and unconditional indications. Finally, we fit a simple linear model (2022–2024) to project future backlog growth under current policies.

**Results:** Out of 613 full HTA reports covering 666 indications, 44% were conditionally approved, 42% unconditionally, and 14% received a negative decision; oncology accounted for ∼40% of all indications. The HTA process (submission to decision) improved considerably, with mean durations nearly halving from 208 days in 2020 to roughly 100 days in 2024. Despite these improvements, the mean time from HTA decision to reimbursement rose from 222 days in 2020 to 461 days in 2024 overall, with conditional decisions taking on average 274 more days than unconditional ones in 2024. Kaplan-Meier analysis showed that by 24 months post-HTA decision, 98.3% of unconditional indications were reimbursed, compared to only 60.1% of conditional indications. Meanwhile, the backlog of unreimbursed indications increased from 47 in 2022 to 146 in 2024, and linear projections suggest it could reach 247 by 2026 under the current system.

**Discussion:** Despite some efficiency gains in the HTA evaluation stage, Romania’s conditional reimbursement pathway remains hampered by tight budgets and administrative hurdles, prolonging patient inaccessibility—particularly in oncology, where timely treatment is critical. Strengthening administrative capacity, diversifying Managed Entry Agreement (MEA) models, and integrating new digital tools could help address these bottlenecks. Without substantial reforms, the backlog will continue growing, limiting patients’ timely access to innovative therapies.

## 1. Introduction

Ensuring timely access to innovative medicines is a fundamental goal of modern healthcare systems. In Romania, drugs seeking reimbursement after approval by the European Medicines Agency (EMA) need to follow a multi-step process to become reimbursed and reach patients. First, they need to undergo a health technology assessment (HTA) by the National Drug Agency (NDA). This scorecard-based Health Technology Assessment (HTA) (Radu et al., 2016) can result in either a denial of reimbursement or a positive decision for reimbursement that can be unconditional or conditional. Unconditional decisions require inclusion in the National Reimbursement List and the publication of Therapeutic Protocols before being reimbursed for the patients. Conditional decisions, by contrast, require an additional step: negotiating a Cost-Volume (CV) or Cost-Volume-Result (CVR) agreement between the Market Authorization Holder (MAH) and the National Health Insurance House (NHIH; the unique national payer).

These Managed Entry Agreements (MEAs), valid for only one year, operate within a limited budget pool approved annually by the Government and must accommodate both new conditional agreements and the annual renewal of existing ones. Between the two, CV agreements are the most widespread. CVR agreements are less frequent and currently limited to new-generation drugs for hepatitis.

Introduced in 2015, the MEA framework for drugs with conditional HTA approval was designed to balance the introduction of innovative therapies with fiscal sustainability (Radu et al., 2023). However, anecdotal evidence suggests that the system has struggled to meet its objectives.

Reports from stakeholders highlight lengthy delays, a lack of predictability, and growing inefficiencies in the reimbursement process. Despite these observations, there has been no systematic, data-driven assessment to quantify the extent of these delays or evaluate their evolution over time. Understanding these issues is critical, particularly as the complexity of conditional agreements continues to grow, placing additional strain on the system and currently preventing access to innovative drugs approved by the EMA.

The challenges in reimbursements are not unique to Romania. Even though the Government and other relevant institutions implemented minor updates to the framework, the country continuously ranks among those with the longest delays between EMA approval and patient access, as highlighted by the European Federation of Pharmaceutical Industries and Associations (EFPIA) W.A.I.T. reports (EFPIA Patients W.A.I.T. Indicator, 2024). While these aggregated metrics shed light on the overall extent of the problem, they do not offer a detailed view of the specific phases in the reimbursement process or the underlying factors driving such delays. A comprehensive evaluation of the Romanian system is needed to identify bottlenecks, particularly in transitioning HTA decisions into actionable reimbursements.

This study provides the first large-scale, systematic analysis of Romania’s reimbursement system. Its objectives are threefold: first, to quantify the magnitude of delays in the reimbursement process over the past decade; second, to assess how these delays have evolved over time, with a specific focus on the differences between conditional and unconditional decisions; and third, to identify trends and highlight the growing backlog of indications awaiting reimbursement. By addressing these objectives, the study aims to provide policymakers with the evidence needed to design more efficient, predictable, and patient-centered reimbursement frameworks.

## 2. Methods

### 2.1. Data eligibility

We included all dossiers submitted from January 1^st^, 2015, and posted on the NDA website by December 31^st^, 2024 (a full 10-years database). Although the Romanian HTA process started in 2014, we excluded dossiers submitted during that initial year. The rationale was threefold: 2014 represented a calibration period where HTA reports were less structured, there was an unusually high negative decision rate, and the conditional reimbursement framework was only implemented starting in 2015. Including 2014 data would not offer a coherent, representative view of the current system. By starting with 2015, we maintain a baseline that accurately reflects the ongoing reimbursement landscape.

Additionally, we excluded dossiers that do not represent the standard reimbursement process, such as those evaluated under “table no. 1” (e.g., new concentrations, pharmaceutical forms, expanded target populations, or different treatment lines) and “table no. 9” (generics or biosimilars of conditionally reimbursed indications). These cases, as well as cost-minimization dossiers, typically entail simpler, faster evaluations and often only require updates to existing Therapeutic Protocols, rather than the full reimbursement pathway. We also excluded HTA reports initiated by the National Health Insurance House to reconfirm existing reimbursements or modify prescription procedures, as they lie outside the standard process for new drugs or indications. By focusing on “full HTA” dossiers—those involving genuinely new drugs or entirely new indications—we ensure a coherent dataset that accurately captures the multifaceted and time-intensive nature of the Romanian reimbursement process.

### 2.2. Data sources and extraction

The main data source was the website of Romania’s National Drug Agency (NDA)—where all publicly available HTA reports submitted by pharmaceutical companies were identified and retrieved. Each retrieved HTA report followed a relatively standardized format. Custom Python scripts—using text normalization and regular expressions—were used to extract information such as the international nonproprietary name (INN), brand name, Anatomical Therapeutic Chemical (ATC) classification code, stated indication(s), HTA submission and decision dates, evaluation track, and final reimbursement decision (unconditional, conditional, or negative).

Data points that could not be automatically extracted were completed manually by referring directly to the HTA reports.

All automatically extracted data were visually inspected to ensure accuracy and consistency. In some cases, certain variables (e.g., HTA decision dates) were not consistently reported by the NDA, particularly before 2020; these were recorded as “Not specified”. Additional Python scripts were used to standardize the formatting and maintain internal consistency.

An “indication” was defined as any distinct subpopulation or treatment scenario evaluated independently by the NDA. A single HTA dossier could generate multiple indications if the NDA issued separate decisions (unconditional, conditional, or negative) for different patient subgroups (e.g., adult vs. pediatric, varying disease severity). This approach allows for a more granular analysis of reimbursement decisions originating from the same dossier.

The initial outcome of an HTA report can be appealed by the MAH. All appeal decisions were manually reviewed and integrated into the original reports, with the date of the appeal decision treated as the final decision date. Cases undergoing appeals were labeled accordingly.

### 2.3. Therapeutic area classification

Therapeutic areas were initially assigned based on the drug’s ATC code as reported in the HTA documents. Each ATC code was mapped to predefined therapeutic areas. A secondary classification method was employed to account for instances where the ATC code was not provided in the HTA report or did not clearly correspond to the specific indication of the drug. Using the OpenAI API (gpt-4o-mini model), we analyzed the stated indication from the HTA report and classified it into one of the 12 categories. All classifications that conflicted with the ATC-based assignment (n=160) were manually reviewed. Following this reconciliation, 123 classifications were ultimately retained as determined by the API-based method.

### 2.4. Determination of reimbursement dates

For all indications with a positive HTA decision (unconditional or conditional), we identified the date of inclusion in the National Reimbursement List and the date of publication for the corresponding Therapeutic Protocol. To accomplish this, all versions of the National Reimbursement List and the List of Therapeutic Protocols (publicly available regulatory documents updated since 2008) were compiled into Excel files, with each version on a separate sheet. An initial Python-based matching process assigned reimbursement dates according to predefined logic rules (e.g., for single-indication drugs, the earliest list entry served as the inclusion date). Complex cases, such as multiple indications requiring protocol amendments, were resolved manually. Two authors independently verified all automated matches and crosschecked their findings; any disagreements were resolved by a third author.

Because reimbursement could occur through either inclusion in the Reimbursement List (with or without protocol changes) or publication of a new Therapeutic Protocol, the final reimbursement date was defined as the later of the two events, providing a consistent measure of when full reimbursement became available. All indications for which a reimbursement date was established were labeled as “reimbursed”.

### 2.5. Key durations

Every indication in the dataset—regardless of reimbursement status—has an HTA submission date, most indications from 2020 onward have an HTA decision date, and reimbursed indications have a reimbursement date. From these data points, we calculated two key time intervals in days:

- Duration of the HTA process, from HTA submission to HTA decision: for all indications.
- Duration from HTA decision to Reimbursement: for reimbursed indications only.

### 2.6. Classification of non-reimbursed and waiting indications

For indications with a positive HTA decision for which we could not identify a valid reimbursement date, we distinguished between “not reimbursed” and “waiting” status based on the HTA submission date. Indications submitted before 2021 were labeled “not reimbursed,” while those submitted from 2021 onward were considered “waiting.” This cutoff reflects both stakeholder feedback and the evolving nature of Romania’s conditional reimbursement pathway. In practice, all “not reimbursed” cases were conditional. Some companies later re-submitted the HTA file with additional evidence or through a different track, occasionally securing an unconditional decision for a narrower patient population. Meanwhile, “waiting” indications— also largely conditional—had typically progressed further (e.g., obtaining population eligibility publication), a key step before negotiation. Informal discussions with industry representatives and the National Health Insurance House (NHIH) confirmed that certain indications have remained in this “waiting” status since 2021.

### 2.7. Descriptive analyses

We summarized the dataset and examined trends in both the HTA and reimbursement processes. This included tabulating indications from the full set of HTA dossiers and categorizing them by therapeutic area, HTA decision type (unconditional, conditional, or negative), and appeal status. We used stacked bar plots (by submission year) to visualize trends in positive HTA decisions and summarized reimbursement outcomes (reimbursed, waiting, or not reimbursed) by submission year, separately for conditional and unconditional indications. For durations, we calculated both the mean (with 95% confidence intervals) and the median (with interquartile ranges). Durations of the HTA process (submission to decision) were analyzed by submission year, while durations from HTA decision to reimbursement were analyzed by reimbursement year, split into conditional vs. unconditional decisions.

We used the HTA submission year for metrics related to the evaluation phase (e.g., number of positive decisions, submission-to-decision duration) to ensure complete coverage, as all indications have a submission date. For post-HTA processes (e.g., decision-to-reimbursement duration), we used the reimbursement year to avoid underestimating recent durations, given that many newer indications remain in “waiting” status. This complementary approach captures both how the system processes incoming HTA dossiers and how effectively it transitions positive decisions into actual reimbursements, minimizing bias in the analysis.

### 2.8. Time-to-reimbursement analysis

To assess time-to-reimbursement, we performed a Kaplan-Meier analysis on indications with a positive HTA decision and a valid decision date, submitted from 2020 onward. We excluded “not reimbursed” indications (final outcome) and retained those classified as “reimbursed” or “waiting.” The time variable extended from the HTA decision date to either the reimbursement date (event) or December 31^st^, 2024 (censor). We stratified by decision type (conditional vs. unconditional) and generated Kaplan-Meier curves to estimate the cumulative probability of reimbursement over time. A log-rank test compared the conditional and unconditional curves, and we reported the percentage of indications reimbursed at 6, 12, and 24 months from the decision date to illustrate disparities.

### 2.9. Backlog size projections

To project the backlog of indications awaiting reimbursement, we defined “backlog” as the number of positive HTA decisions still not reimbursed by the end of the year being analyzed.

From 2022 to 2024, the backlog increased at a relatively linear pace. Using these three data points, we fit a simple linear regression model of the form *waiting* = *β*_0_ + *β*_1_ * *year* to predict backlog sizes for 2025 and 2026, including 95% confidence intervals. This model assumes no major changes to reimbursement policies, budgets, or the influx of new positive decisions. Its simplicity mirrors the observed linear growth and helps minimize overfitting.

### 2.10. Software used

All data processing and cleaning were conducted using Python (v3.11). Statistical analyses and visualizations were performed using R (v4.4.0) within the RStudio environment (v2024.04.1).

### 2.11. Ethical considerations

All data used in this study are publicly available and do not contain patient-level information. Therefore, no ethical approval was deemed necessary.

## 3. Results

We identified 613 unique HTA reports meeting the inclusion criteria (full HTA). These correspond to 666 distinct indications, as defined by the NDA. The main characteristics of the indications analyzed are presented in Table 1. As noted in the table, a considerable proportion (40%) of the indications submitted by pharmaceutical companies for full HTA evaluation belong to the field of oncology. Except for the area of autoimmune & inflammatory diseases (accounting for 15%), the split between other therapeutic areas is relatively uniform, ranging between 2% and 9%. In terms of the HTA outcome, 14% of indications received a negative decision, while the remaining are almost equally split between conditional (44%) and unconditional (42%). Fewer than 5% of all indications from full HTA evaluation have gone through the appeal process, which reflects the high acceptability of the initial HTA outcome.

**Table 1.** Descriptive characteristics of included indications (N=666).

| Characteristic | n (%) |
| --- | --- |
| **Therapeutic area** |  |
| oncology | 267 (40%) |
| autoimmune & inflammatory diseases | 98 (15%) |
| hematology | 57 (8.6%) |
| neurology & psychiatry | 41 (6.2%) |
| endocrinology & metabolism | 39 (5.9%) |
| other | 34 (5.1%) |
| infectious diseases | 33 (5.0%) |
| cardiovascular diseases | 31 (4.7%) |
| respiratory diseases | 21 (3.2%) |
| dermatology | 18 (2.7%) |
| gastroenterology & hepatology | 14 (2.1%) |
| renal & nephrology | 13 (2.0%) |
| **HTA decision** |  |
| conditional | 294 (44%) |
| unconditional | 281 (42%) |
| negative | 91 (14%) |
| **Appeal** |  |
| no | 636 (95%) |
| yes | 30 (4.5%) |

Full HTA submissions between 2015 and 2020 resulted in a growing number of indications with a positive decision each year, followed by a plateau in more recent years (Figure 1). While for 2024 it might appear there are fewer positive decisions, this likely reflects incomplete processing of dossiers submitted in 2024 by the NDA at the study’s cutoff date (December 31^st^, 2024). As shown in the figure by the percentages in parentheses, the proportion of conditional decisions by year of HTA submission has registered a considerable increase versus unconditional decisions along the years. The split between conditional and unconditional decisions has remained at a steady 2:1 ratio respectively for dossiers submitted starting with 2022. The rising proportion of conditional decisions, particularly after 2022, signals growing systemic complexity and may contribute to the increasing backlog observed in recent years.

**Figure 1.**
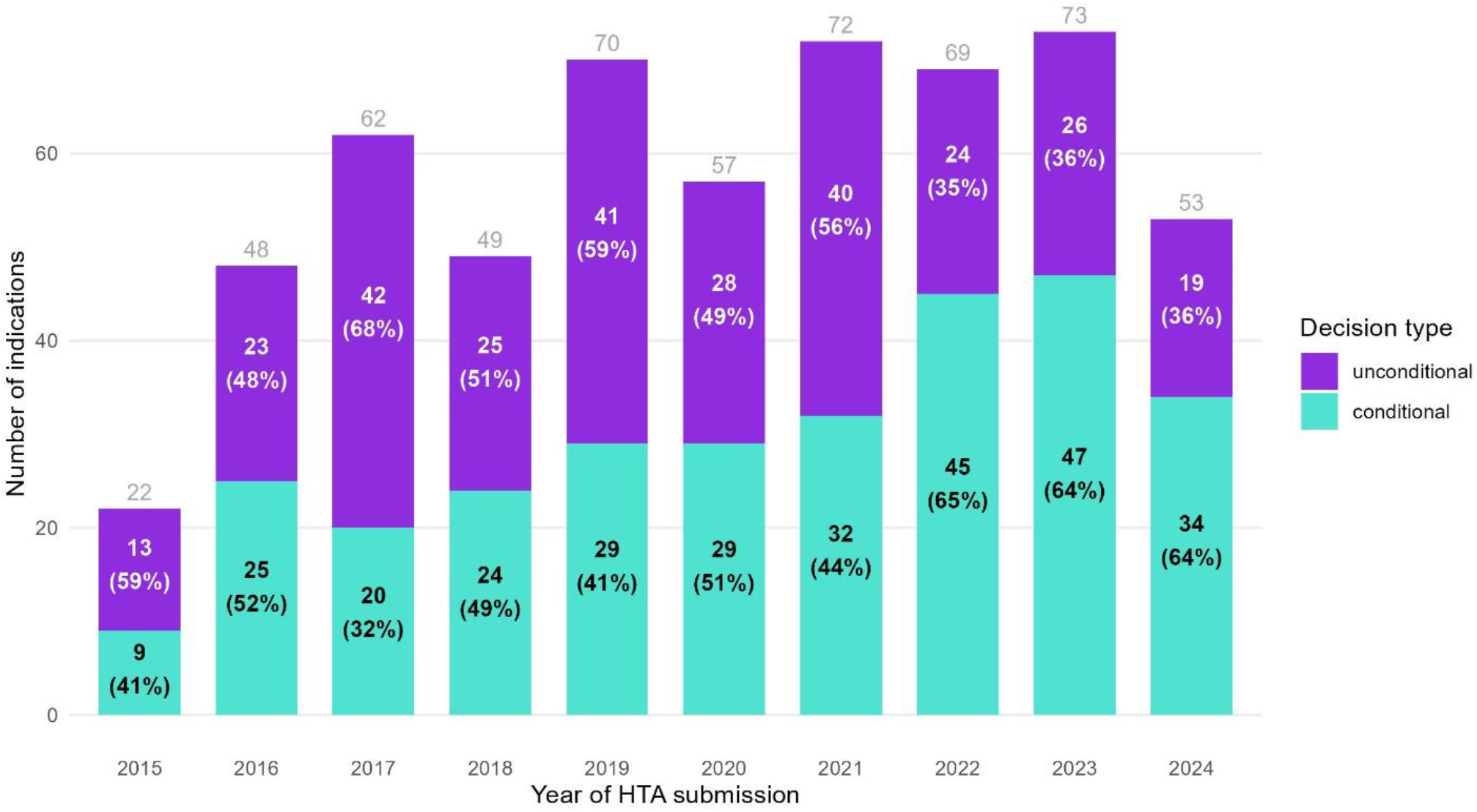
Indications with a positive HTA decision, split by decision type and year of HTA submission

Figure 2 enriches the perspective on the distribution between conditional and unconditional decisions by reimbursement status (exact numbers in Supplemental Table 1). Indications with a conditional decision disproportionately contribute to the backlog, as they are more likely to remain either not reimbursed or waiting at the study’s cutoff date. 18.8% of the conditional decisions from dossiers submitted in 2021 were waiting for reimbursement, compared to only 2.5% of the unconditional ones. For dossiers submitted in 2022, 66.7% of the conditional decisions were waiting to be reimbursed at the study’s cutoff date, compared to only 4.2% of the unconditional decisions. For dossiers submitted in 2023, the percentages converge, and for 2024, conditional decisions reach 100% pending reimbursement. These findings underscore the delays faced by conditional indications, which are particularly pronounced in recent submission years. The growing disparity between conditional and unconditional reimbursement outcomes highlights structural inefficiencies in the current reimbursement framework.

**Figure 2.**
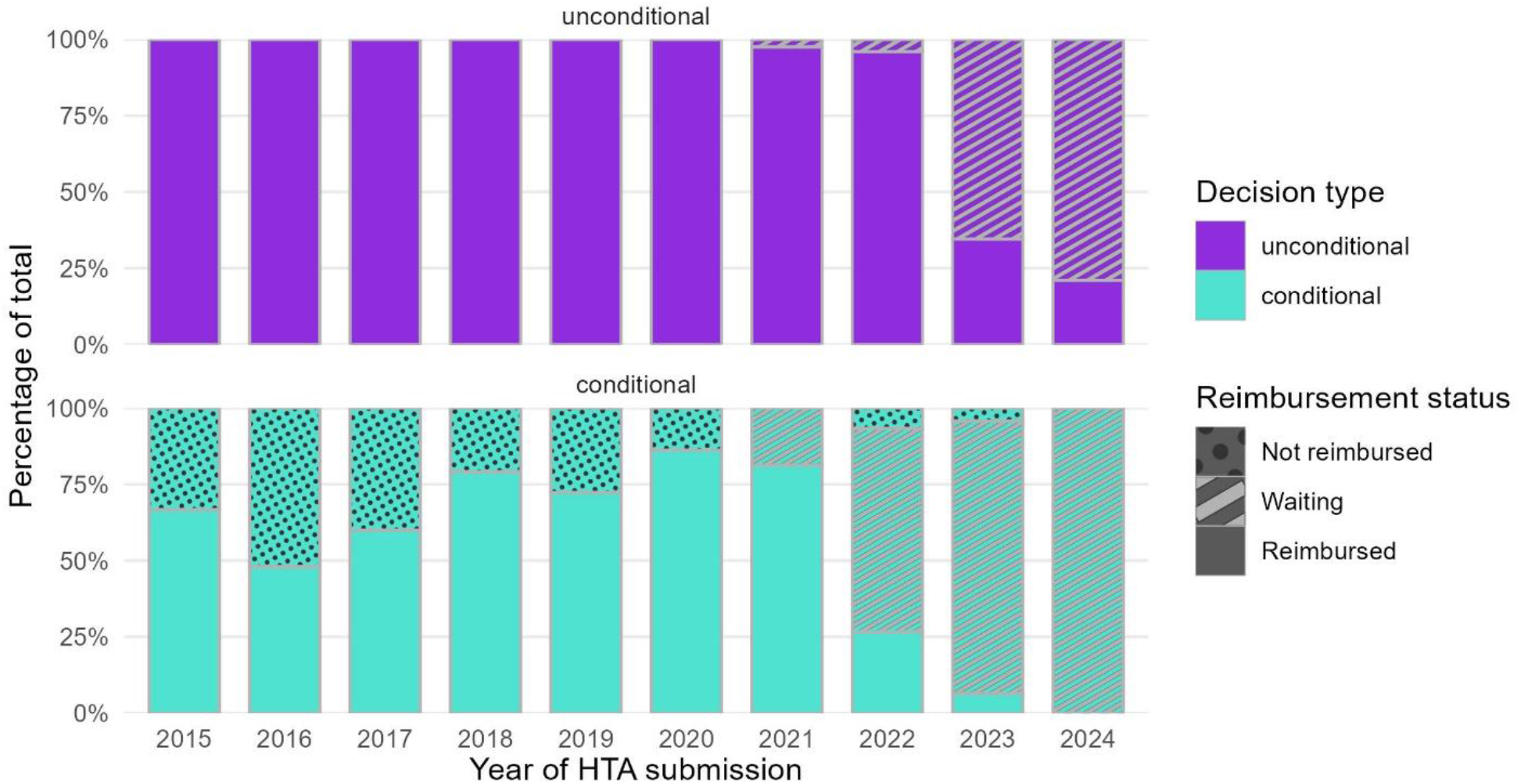
Distribution of positive HTA decisions by decision type and year of HTA submission

An abrupt shift is observed in the reimbursement pattern of conditional decisions (predominantly CV agreements) starting with HTA files submitted in 2022. The proportion of these decisions achieving reimbursement declined sharply from over 80% in previous years to 27% in 2022. This reduction is likely driven by budgetary limitations but is also influenced by administrative hurdles.

In terms of durations, we were able to analyze indications from dossiers submitted starting 2020, as previous dossiers were inconsistent in reporting the HTA decision date. As seen in Table 2, the HTA process—performed entirely by the NDA—has become much quicker in 2024 compared to 2020, with a halving of the mean and median times between these years to roughly three months. This improvement may reflect greater efficiency within the NDA, confirming that the growing delays in reimbursement are not caused by the HTA process itself.

**Table 2.** Mean and median values for the HTA process duration (days), split by submission year.

| Submission<br>year | Overall |  |
| --- | --- | --- |
|  | Mean [95% CI] | Median [IQR] |
| 2020 | 208 [184–232] | 200 [161–246] |
| 2021 | 259 [236–282] | 235 [194–296] |
| 2022 | 178 [158–197] | 180 [121–209] |
| 2023 | 120 [112–128] | 118 [96–135] |
| 2024 | 94 [86–103] | 94 [80–106] |

In contrast, the overall duration between the HTA decision and reimbursement has increased steadily when examined retrospectively (Supplemental Table 2), reflecting persistent inefficiencies in transitioning positive HTA decisions into reimbursed indications. From a mean duration of 222 days for 2020 reimbursements, indications reimbursed in 2024 have had a mean duration from their HTA decision of 461 days, with median durations following very closely.

These steps depend on the available budget for negotiations of CV agreements by the NHIH and on the periodicity of Reimbursement List updates.

When stratified by decision type (Figure 3), both conditional and unconditional indications show increased durations over time; however, conditional reimbursements consistently exhibit significantly longer delays. The gap is widening considerably for 2024 reimbursements, where conditional indications take on average 274 days longer from HTA decision to reimbursement than unconditional ones. These findings emphasize that while HTA evaluations have become faster, post-HTA reimbursement processes remain a significant bottleneck, particularly for conditional indications.

**Figure 3.**
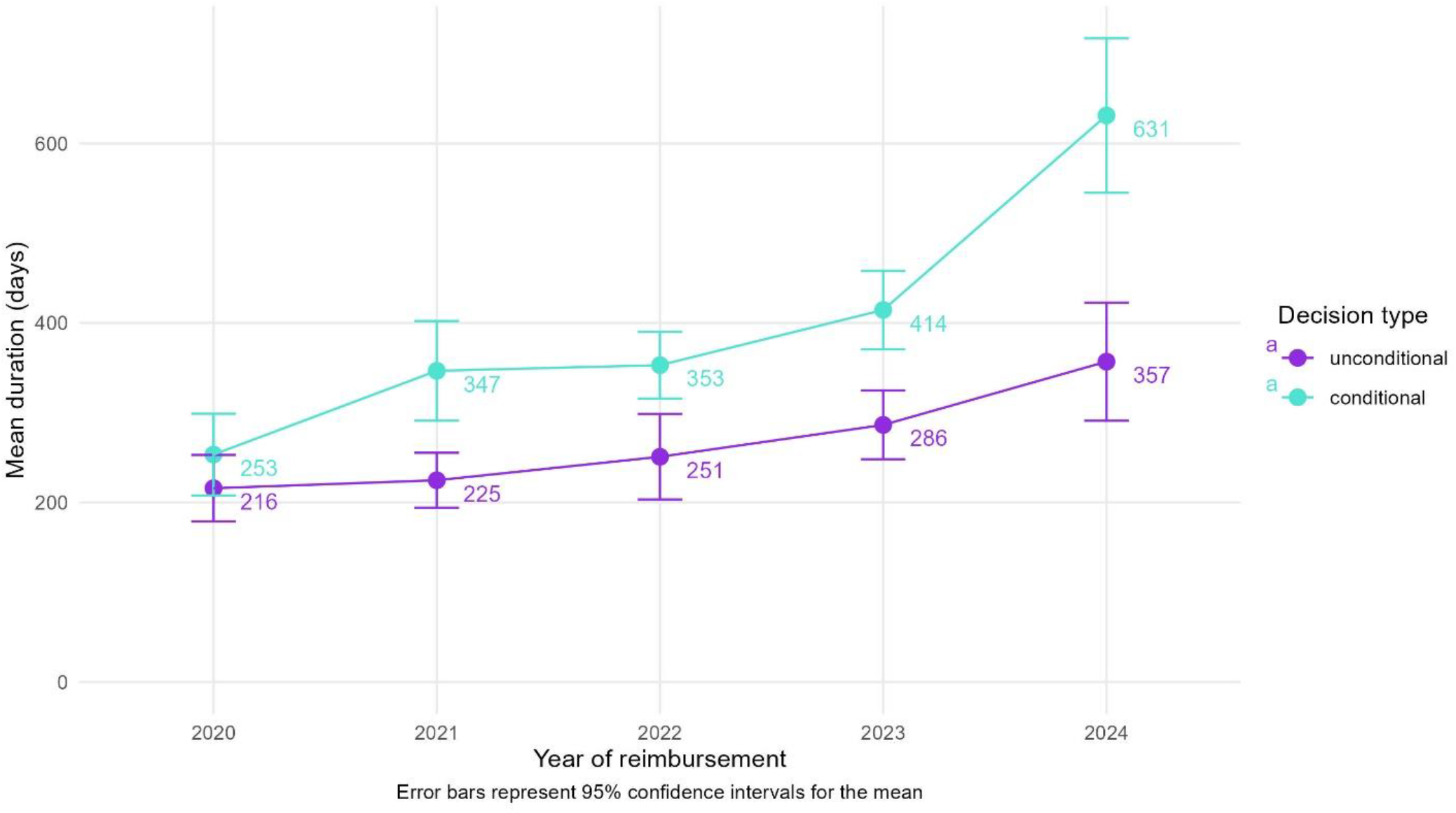
Mean durations between the HTA decision and Reimbursement, split by decision type and year of reimbursement

To explore this further, we performed a Kaplan-Meier survival analysis. This analysis included 300 indications with a positive HTA decision and valid decision dates (submissions from 2020 onward), of which 128 (42.7%) were unconditional and 172 (57.3%) were conditional. The cumulative probabilities of being reimbursed are shown in Figure 4. The reimbursement probability curves indicate significantly longer time-to-reimbursement for conditional indications compared to unconditional indications (log-rank test, p < 0.0001).

**Figure 4.**
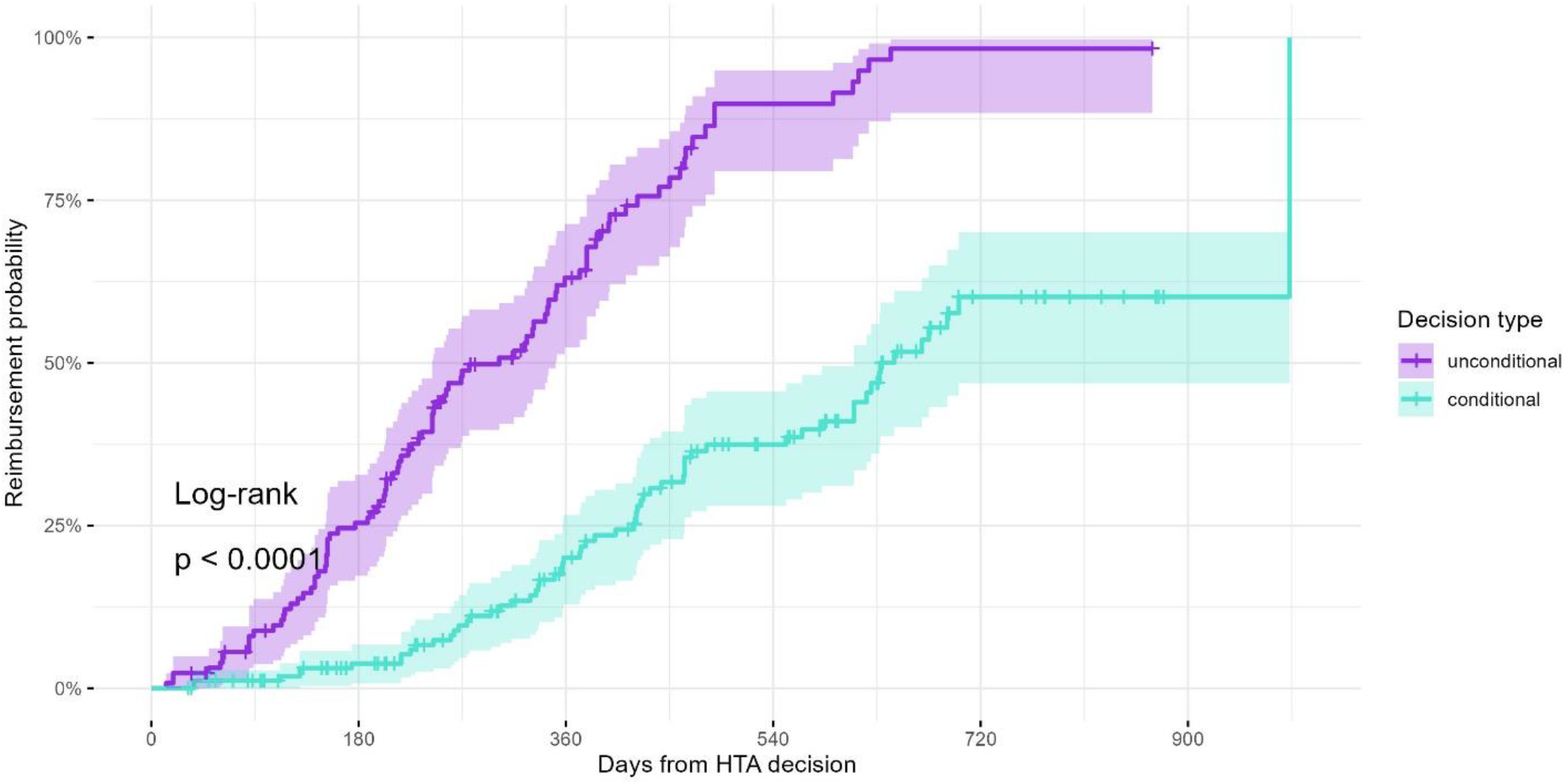
Reimbursement probability curves for all indications with a positive decision from HTA dossiers submitted starting 2020

At 6 months following the HTA decision, 25.4% of unconditional indications were reimbursed, compared to only 3.8% of conditional indications. By 12 months, 63.1% of unconditional indications were reimbursed, while only 20.1% of conditional indications had been reimbursed. At 24 months, nearly all unconditional indications (98.3%) had been reimbursed, compared to only 60.1% of conditional indications.

These results demonstrate a substantial and statistically significant disparity in time-to-reimbursement between conditional and unconditional indications, highlighting the inefficiencies faced by conditional pathways. The divergence between the survival curves suggests that conditional indications face not only slower but also less predictable reimbursement processes.

This disparity likely reflects administrative burdens, negotiation complexities, and budgetary constraints associated with conditional agreements.

The backlog of indications waiting for reimbursement grew from 47 at the end of 2022 to 103 by the close of 2023, reaching 146 by the end of 2024 (Figure 5). Based on the linear model fitted to data from 2022 to 2024, the backlog is projected to grow to 198 indications by the end of 2025 (95% CI: 95–301) and 247 (95% CI: 99–395) indications by the end of 2026.

**Figure 5.**
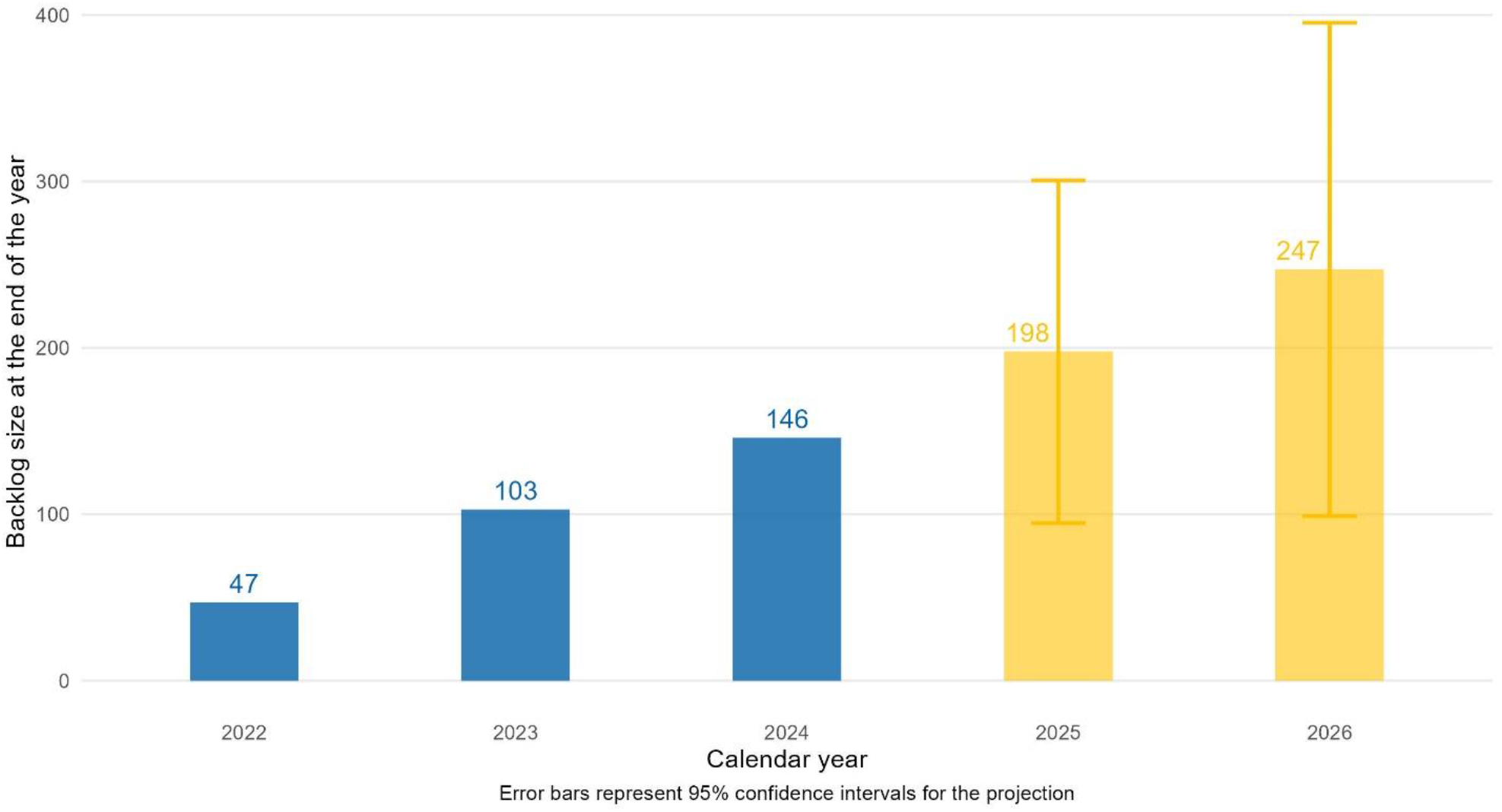
Year-end backlog of indications waiting for reimbursement (blue = actual data for 2022-2024, orange = projected data for 2025-2026)

These findings highlight a growing gridlock in the reimbursement system, where the capacity to reimburse new indications has remained largely static while the number of conditional HTA decisions continues to rise. The static nature of the system’s capacity to handle conditional agreements exacerbates the backlog, particularly for innovative therapies that require additional negotiation and funding under conditional reimbursement pathways.

## 4. Discussion

### 4.1. Interpretation of findings

This study reveals a critical bottleneck in Romania’s reimbursement system for innovative medicines, characterized by significant delays and a growing backlog of indications awaiting reimbursement. Conditional HTA decisions, which require more complex negotiations and administrative oversight, disproportionately contribute to these delays. Although administrative hurdles remain an obstacle, the primary issue is insufficient funding: the volume of conditional decisions and the associated financial requirements for implementing CV agreements exceed the budget allocated by the Government to the NHIH.

The Kaplan-Meier survival analysis and duration statistics reveal pronounced disparities: conditional indications face slower and less consistent reimbursement than their unconditional counterparts. This outcome appears counterintuitive, given that CV agreements were introduced as a mechanism to facilitate faster and broader access through negotiations between pharmaceutical companies and the NHIH. For all drugs reimbursed unconditionally, companies must pay a mandatory fixed 25% tax (“clawback”). For conditionally (CV) reimbursed drugs, companies pay a variable payback rate, depending on the percentage of eligible population being treated; in this case, the legislation imposes a payback grid ranging from 25% to 70% (Government of Romania, 2011). Furthermore, the budget for new CV agreements is separate from the one for unconditional reimbursements and has been capped from 2018 through the fourth quarter of 2024.

Although CV agreements were intended to manage the financial uncertainty associated with innovative therapies, the capped yearly budget—combined with the healthcare system’s limited capacity to track and reconcile expenditures with savings—has ultimately led to a slowdown in expanding the Reimbursement List. These delays have far reaching implications as they result in prolonged inaccessibility to innovative and potentially life-saving treatments, particularly in high-impact therapeutic areas such as oncology and rare diseases.

Our findings align with prior research showing Romania’s prolonged delays in access to new medicines relative to other European countries (EFPIA Patients W.A.I.T. Indicator, 2024).

EFPIA’s WAIT reports have consistently ranked Romania among the countries with the longest delays from EMA approval to patient access. Moreover, Romania also registers one of the lowest percentages of medicines patients have access to in the country compared to the number of medicines approved by the EMA. Our study adds critical nuance by deconstructing delays into specific phases of the reimbursement process and quantifies the backlog of drugs waiting for reimbursement.

While the HTA process itself has become faster, with durations halving from 2022 to 2024, this improvement has been eclipsed by worsening post-HTA delays. The mean duration from HTA decision to reimbursement increased from 222 days for 2020 reimbursements to 461 days for 2024 reimbursements. The backlog of unreimbursed indications illustrates the consequences of these delays. Our data show that the backlog grew from 47 in 2022 to 146 in 2024, with linear projections estimating further growth to 247 indications by 2026 under the status quo. Behind these numbers lies a stark human impact: delayed access to essential treatments, particularly for patients with severe and rapidly progressing conditions such as cancer. With oncology accounting for 40% of the HTA dossiers from the past decade, this reality cannot be overstated. For many patients, these delays are critical and, in some cases, untenable.

### 4.2. Policy recommendations

The findings underscore the urgent need for systemic reforms to Romania’s reimbursement framework. The dominance of conditional HTA decisions reflects both the increasing complexity of innovative therapies and the need for a more flexible MEA framework. Conditional reimbursements require extensive negotiation and monitoring, yet the system remains constrained by static administrative capacity and a limited budget pool. This structural mismatch between growing demand and stagnant capacity exacerbates delays and disproportionately impacts patients requiring access to innovative therapies.

Addressing these challenges necessitates a multifaceted approach. First, early involvement of all relevant stakeholders in horizon-scanning initiatives is critical. This should involve coordinated efforts among the Ministry of Health, Ministry of Finance, Ministry of Economy, NHIH, NDA, and patient associations—especially in view of the forthcoming EU Pharmaceutical legislation— to adapt financing and reimbursement policies to better align with patient needs and the evolving profiles of new EMA-approved therapies. In parallel, the reimbursement process itself must be renewed and integrated to ensure that each segment—ranging from HTA evaluation and MEA negotiation to the publication of the Reimbursement List and Therapeutic Protocols—operates with reasonable and predictable waiting times. Digitalization efforts spearheaded by the Ministry of Health could further enhance efficiency and transparency. Moreover, establishing a predictable budget for MEA negotiations that can dynamically adjust to real-world evidence on the usage of new therapies is imperative. Expanding the MEA portfolio should aim to combine different mechanisms addressing either clinical or financial uncertainty, while providing simpler and more efficient governance using new technology to free up administrative resources. Examples of additional MEAs could encompass both simpler mechanisms—such as upfront discounts, price caps, utilization caps—and more advanced performance- or outcome-based agreements could offer much-needed flexibility. However, implementing more complex MEAs necessitates more human and IT resources (Wenzl and Chapman, 2019; Ferrario et al., 2017). Regardless of the MEA type, improving administrative capacity through both personnel and technology will be critical to streamline governance and reduce delays. Without these comprehensive improvements, the system risks further delays, perpetuating inequitable access and allowing the backlog of indications to continue growing, ultimately jeopardizing timely patient access to innovative therapies.

### 4.3. Strengths and limitations

This study is the first large-scale analysis of reimbursement timelines in Romania, offering robust evidence on systemic inefficiencies. By combining manual and automated data extraction methods, the dataset integrates all publicly available HTA reports since 2015, providing high granularity and accuracy. The use of multiple analytical approaches, including Kaplan-Meier survival analysis and backlog projections, strengthens the reliability of the findings.

The following limitations are worth mentioning. First, recent submissions classified as “waiting” may not ultimately pursue reimbursement, potentially leading to an overestimation of the backlog. Second, missing HTA decision dates, particularly for dossiers submitted before 2020, limited our ability to analyze earlier trends comprehensively. Third, minor adjustments to the HTA framework during the study period were not explicitly accounted for in the statistical models. Finally, the assumption of a status quo for backlog projections may not fully reflect potential policy changes or adjustments in system capacity.

### 4.4. Future directions

This study lays a foundation for future research aimed at improving Romania’s reimbursement framework. Future studies should explore alternative MEA designs tailored to the Romanian context, such as adaptive risk-sharing agreements or performance-based reimbursement models. Simulation models could be employed to assess the feasibility and impact of such agreements on reducing delays and backlog growth. Additionally, ongoing monitoring of reimbursement timelines and outcomes will be critical for evaluating the effectiveness of policy interventions.

Regular audits of the negotiation and reimbursement processes could identify bottlenecks and inform targeted improvements.

In parallel, the new Regulation on health technology assessment from the EU (“EU HTA”), which took effect in January 2025, strives to harmonize HTA processes across EU member states and enhance timely patient access to innovative therapies. The reforms proposed in our study are complementary to this initiative, offering an opportunity for Romania to align its national procedures with broader EU objectives and further improve reimbursement efficiency and timelines.

## 5. Conclusion

This study offers the first comprehensive assessment of Romania’s drug reimbursement system. Although the framework represented a significant advance at its inception, the rapid emergence of new technologies and challenges in resource management have gradually led to delays in patient access and a growing backlog, especially for indications receiving conditional HTA decisions. These systemic issues not only strain administrative processes but also have serious implications for patients, particularly in oncology, where timely access to treatment is critical. Since the existing cost-volume mechanism alone fails to adequately manage financial risks—and the annual drug budget is neither based on horizon scanning nor supported by real-world evidence—the backlog is expected to continue its alarming rise without substantial reforms.

Policymakers must therefore enact meaningful changes to both the MEA framework and funding mechanisms, ensuring a more sustainable and equitable reimbursement environment that better serves patient needs.

## Data Availability

All data produced in the present study are available from public sources.

## Author contributions

CR: conceptualization, methodology, data acquisition, curation, formal analysis, visualization, and drafting of the manuscript. DES: data acquisition and drafting of the manuscript. NDC: conceptualization, supervision, data curation, and drafting of the manuscript.

## Acknowledgements

The authors are grateful to Petter Foss, Kawitha Helme, and Alexandru Diniţă for their feedback on the final version of this manuscript.

## References

Radu, C.P., Chiriac, N.D. and Pravat, A.M. (2016) ‘The Development of the Romanian Scorecard HTA System’, Value Health Reg Issues, 10, pp. 41–47. doi:10.1016/j.vhri.2016.07.006

Radu, C.P., Drăgoi, L., Udroiu, M.A., Pană, B.C. and Iliescu, M.C. (2023) ‘The outcomes of managed entry agreements in Romania from 2015 to 2022’, Farmacia, 71(6), p. 1316. doi:10.31925/farmacia.2023.6.23

EFPIA Patients W.A.I.T. Indicator (2024) EFPIA Patient W.A.I.T. Indicator 2024 Survey. Available at: https://efpia.eu/media/vtapbere/efpia-patient-wait-indicator-2024.pdf x(Accessed: 6 February 2025).

Wenzl, M. and Chapman, S. (2019) ‘Performance-based managed entry agreements for new medicines in OECD countries and EU member states: How they work and possible improvements going forward’, OECD Health Working Papers, No. 115, OECD Publishing, Paris. doi:10.1787/6e5e4c0f-en

Ferrario, A., Araja, D., Bochenek, T. et al. (2017) ‘The Implementation of Managed Entry Agreements in Central and Eastern Europe: Findings and Implications’, Pharmacoeconomics, 35(12), pp. 1271–1285. doi:10.1007/s40273-017-0559-4

Government of Romania (2011) Ordonanţă de urgenţă nr. 77/2011 privind stabilirea unor contribuţii pentru finanţarea unor cheltuieli în domeniul sănătăţii [Emergency Ordinance No. 77/2011 on establishing certain contributions to finance healthcare expenditures]. Available at: https://legislatie.just.ro/Public/DetaliiDocument/131707 x(Accessed: 6 February 2025).

