## Supplemental materials for "Delayed Access to Innovative Medicines in Romania: A Comprehensive Analysis of the Reimbursement Processes (2015-2024)"

[1]Novartis Pharma Services, Bucharest, Romania.

**Supplemental material**

**Supplemental Table 1.** Detailed trends for indications with a positive HTA decision: crosstabulation of decision type and reimbursement status

| | | Reimbursed | | Waiting | | Not reimbursed | | |
|---|---|---|---|---|---|---|---|---|
| **Submission year** | **Decision type** | **n** | **%** | **n** | **%** | **n** | **%** | **Total n** |
| 2015 | unconditional | 13 | 100.0 | 0 | 0.0 | 0 | 0.0 | 13 |
| 2015 | conditional | 6 | 66.7 | 0 | 0.0 | 3 | 33.3 | 9 |
| 2016 | unconditional | 23 | 100.0 | 0 | 0.0 | 0 | 0.0 | 23 |
| 2016 | conditional | 12 | 48.0 | 0 | 0.0 | 13 | 52.0 | 25 |
| 2017 | unconditional | 42 | 100.0 | 0 | 0.0 | 0 | 0.0 | 42 |
| 2017 | conditional | 12 | 60.0 | 0 | 0.0 | 8 | 40.0 | 20 |
| 2018 | unconditional | 25 | 100.0 | 0 | 0.0 | 0 | 0.0 | 25 |
| 2018 | conditional | 19 | 79.2 | 0 | 0.0 | 5 | 20.8 | 24 |
| 2019 | unconditional | 41 | 100.0 | 0 | 0.0 | 0 | 0.0 | 41 |
| 2019 | conditional | 21 | 72.4 | 0 | 0.0 | 8 | 27.6 | 29 |
| 2020 | unconditional | 28 | 100.0 | 0 | 0.0 | 0 | 0.0 | 28 |
| 2020 | conditional | 25 | 86.2 | 0 | 0.0 | 4 | 13.8 | 29 |
| 2021 | unconditional | 39 | 97.5 | 1 | 2.5 | 0 | 0.0 | 40 |
| 2021 | conditional | 26 | 81.2 | 6 | 18.8 | 0 | 0.0 | 32 |
| 2022 | unconditional | 23 | 95.8 | 1 | 4.2 | 0 | 0.0 | 24 |
| 2022 | conditional | 12 | 26.7 | 30 | 66.7 | 3 | 6.7 | 45 |

| Submission year | Decision type | Reimbursed n | Reimbursed % | Waiting n | Waiting % | Not reimbursed n | Not reimbursed % | Total n |
|---|---|---|---|---|---|---|---|---|
| 2023 | unconditional | 9 | 34.6 | 17 | 65.4 | 0 | 0.0 | 26 |
| 2023 | conditional | 3 | 6.4 | 42 | 89.4 | 2 | 4.3 | 47 |
| 2024 | unconditional | 4 | 21.1 | 15 | 78.9 | 0 | 0.0 | 19 |
| 2024 | conditional | 0 | 0.0 | 34 | 100.0 | 0 | 0.0 | 34 |

*Note: all percentages (%) are row percentages and represent the proportion of each reimbursement status within the total HTA decisions for each inclusion type and submission year.*

**Supplemental Table 2.** Mean and median values for the HTA decision to Reimbursement duration (days), split by reimbursement year

| Reimbursement year | Overall | | Unconditional | | Conditional | |
|---|---|---|---|---|---|---|
| | Mean [95% CI] | Median [IQR] | Mean [95% CI] | Median [IQR] | Mean [95% CI] | Median [IQR] |
| 2020 | 222 [190–254] | 228 [182–272] | 216 [179–253] | 225 [180–272] | 253 [208–299] | 238 [230–268] |
| 2021 | 276 [244–309] | 273 [190–330] | 225 [194–255] | 235 [155–299] | 347 [291–402] | 331 [262–415] |
| 2022 | 298 [265–331] | 286 [193–378] | 251 [203–298] | 214 [151–340] | 353 [316–390] | 358 [290–421] |
| 2023 | 318 [281–356] | 322 [244–389] | 286 [248–325] | 262 [244–340] | 414 [371–458] | 430 [386–440] |
| 2024 | 461 [396–525] | 470 [324–623] | 357 [291–423] | 393 [198–473] | 631 [545–717] | 632 [582–675] |
